# SARS-CoV-2 antigen rapid diagnostic test enhanced with silver amplification technology

**DOI:** 10.1101/2021.01.27.21250659

**Authors:** Kei Miyakawa, Rikako Funabashi, Yutaro Yamaoka, Sundararaj Stanleyraj Jeremiah, Junichi Katada, Atsuhiko Wada, Toshiki Takei, Kohei Shimizu, Hiroki Ozawa, Chiharu Kawakami, Shuzo Usuku, Nobuko Tanaka, Etsuko Yamazaki, Hideaki Shimizu, Nobuhiko Okabe, Hideki Hasegawa, Akihide Ryo

**Author notes:** **To whom correspondence should be addressed: Akihide Ryo, M.D., Ph.D.**, Department of Microbiology, Yokohama City University School of Medicine 3-9 Fuku-ura, Kanazawa, Yokohama, Kanagawa 236-0004, Japan.

## Abstract

Rapid diagnosis of COVID-19 is essential for instituting measures to prevent viral spread. SARS-CoV-2 antigen rapid diagnostic test (Ag-RDT) based on lateral flow immunochromatography assay (LFIA) principle can visually indicate the presence of SARS-CoV-2 antigens as a band. Ag-RDT is clinically promising as a point-of-care testing because it can give results in a short time without the need for special equipment. Although various antigen capture LFIAs are now available for rapid diagnosis for SARS-CoV-2 infection, they face the problems of low sensitivity. We have previously developed highly specific monoclonal antibodies (mAb) against SARS-CoV-2 nucleocapsid protein (NP) and in this study, we have employed these mAbs to develop a new LFIA that can detect SARS-CoV-2 NP in nasopharyngeal swab samples with higher sensitivity by combining them with silver amplification technology. We also compared the performance of our Ag-RDT against the commercially available Ag-RDTs using clinical samples to find that our newly developed LFIA performed best among tested, highlighting the superiority of silver amplification technology.

## Introduction

The pandemic of COVID-19 caused by SARS-CoV-2 has brought in serious health threats, adverse economic impacts, and has modified lifestyles in a global scale. Until there is universal access to effective vaccines, the only fail proof way to prevent the spread of the infection is by rapid detection and quarantine of SARS-CoV-2-infected individuals and their contacts (Sun et al., 2020). In this regard, nucleic acid amplification tests (NAAT), such as reverse transcription quantitative polymerase chain reaction (RT-qPCR), is the most sensitive and reliable test that is currently being used (Sethuraman et al., 2020). However, being a centralized test requiring specialized equipment and facility, NAAT might not be readily available in several remote areas. Also, NAAT requires technical processes such as viral RNA extraction from clinical specimens which require technical expertise and prolong the turn-around-time.

On the other hand, SARS-CoV-2 antigen rapid diagnostic test (Ag-RDT) can be a point-of-care test (POCT) providing results within a 30 minutes of sample collection without the need for special equipment and trained personnel. Most SARS-CoV-2 Ag-RDT kits utilize the antigen capture sandwich principle employing a simple lateral flow immunochromatography assay (LFIA) platform. The antigen in the nasopharyngeal swab sample dropped into the device, slowly flows on the cellulose membrane strip together with the colloidal gold-labeled antibody, and when captured by the membrane-bound capture antibody, it develops color and appears as a distinct band (Dinnes et al., 2020; Yamayoshi et al., 2020). However, problems such as subjective visual misidentification, lower sensitivity and occurrence of false positives are the drawbacks of Ag-RDTs.

To overcome these problems, we recently developed a pair of highly specific monoclonal antibodies (mAbs) against the SARS-CoV-2 nucleocapsid protein (NP). These mAbs exclusively recognize the NP of SARS-CoV-2, but not that of SARS-CoV or other coronaviruses. Using these mAbs we developed a highly specific prototype LFIA and improved its sensitivity by enhancing the positive band signal with the Silver halide photography technology. We thus developed the YCU-FF LFIA, an improved Ag-RDT that can test positive for as low as 56 copies/µl of SARS-CoV-2 in test samples, making it more sensitive than the commercially available LFIA based Ag-RDT kits. YCU-FF LFIA could enable rapid diagnosis of SARS-CoV-2 infection with better precision in medical facilities and field settings.

## Results

We have previously reported the production of highly specific mAbs against SARS-CoV-2 NP protein (Yamaoka et al., 2020). To generate LFIA-based SARS-CoV-2 Ag-RDT, we utilized silver halide photography technology for output signal amplification (Broger et al., 2019; Mitamura et al., 2013; Wada et al., 2011; Welch et al., 2017) (Figure 1A). In this technology, silver ions are allowed to adhere to the surface of gold nanoparticles (approx. 0.05 µm in diameter) which allows the electrons from the solution to reduce the silver atoms. This causes further attachment and reduction within a period of 30 seconds, resulting in size enhancement (approx. 5-10 µm in diameter) to provide 1000-fold improvement in visibility (Figure 1B, C). By combining the above technology with specific mAbs, the YCU-FF LFIA provides superior visual detection of antigen-antibody complexes (Supplementary Figure S1).

**Figure 1.**
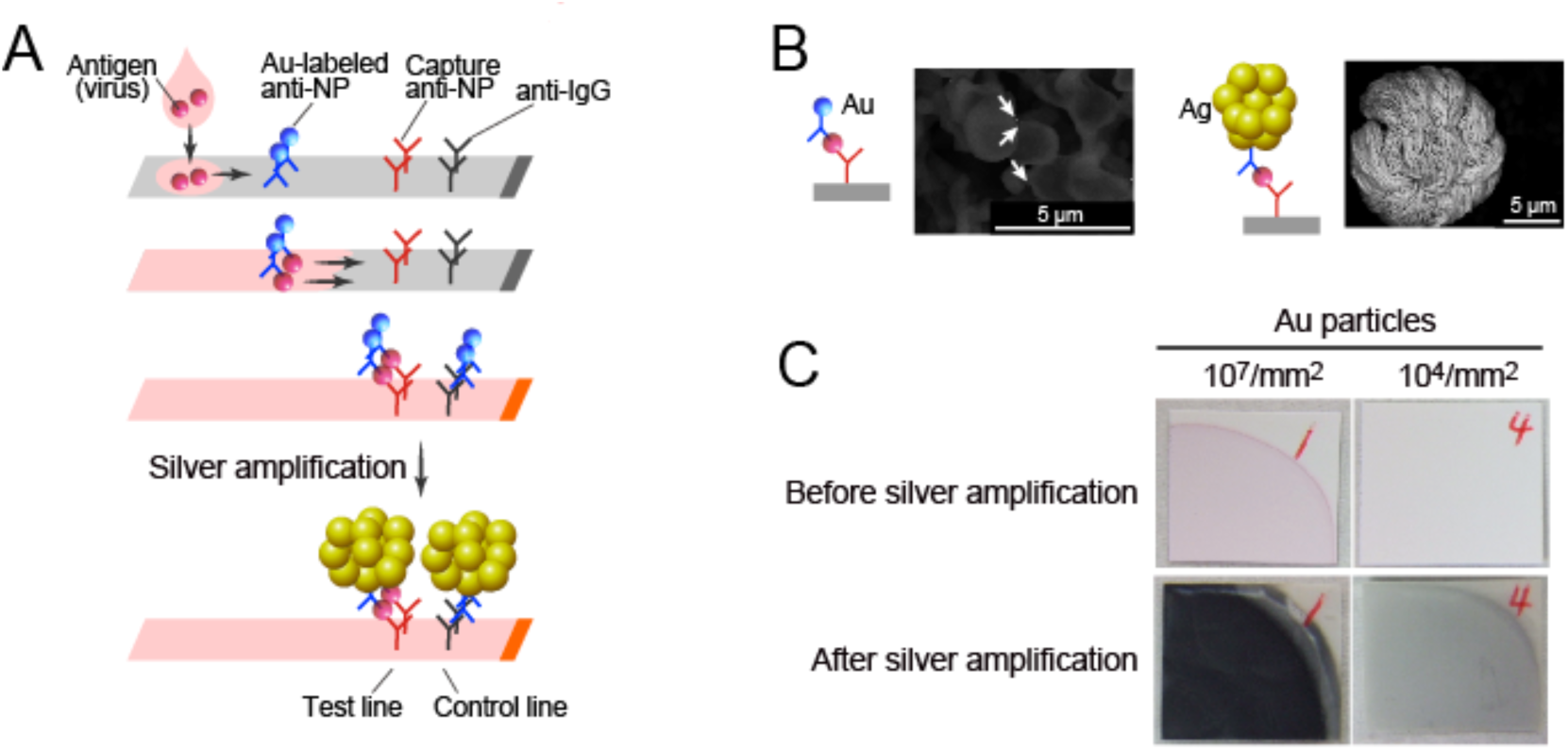
SARS-CoV-2 Ag-RDT with silver amplification technology. **(A)** Schematic diagram of lateral flow immunoassay with silver amplification technology. The antigen in the sample dropped into the device flows on the cellulose membrane together with the colloidal gold-labeled anti-SARS-CoV-2 NP antibody, and when captured by the membrane-immobilized capture antibody, it develops color and appears as a single band. Adherence of silver ions to the surface of a catalytic gold nanoparticle causes electrons to reduce the silver atoms, leading to the size enhancement followed by 1000-fold improvement in visibility. **(B, C)** Differences in SEM images (B) and naked eye visualized bands (C) with and without silver amplification.

YCU-FF LFIA showed no cross-reactivity to related human coronaviruses or other respiratory viruses, indicating its high specificity (Figure 2A). Even the SARS-CoV NP antigen, which has the highest homology with SARS-CoV-2, was tested negative, as our mAbs did not bind SARS-CoV NP (Yamaoka et al., 2020). We then tested if YCU-FF LFIA identified authentic SARS-CoV-2 viruses from culture, and we found that YCU-FF LFIA tested positive for the strain from 56 copies/µl (Figure 2B). This was tested in parallel experiments with commercially available LFIA-based Ag-RDTs; Abbott Panbio COVID-19 Ag Rapid Test, Roche SARS-CoV-2 Rapid Antigen Test, SD Biosensor Standard Q COVID-19 Ag, and Fujirebio Espline SARS-CoV-2; to reveal that YCU-FF LFIA had a superior performance with respect to the minimum limit of detection (LOD) compared to the others (Figure 2B).

**Figure 2.**
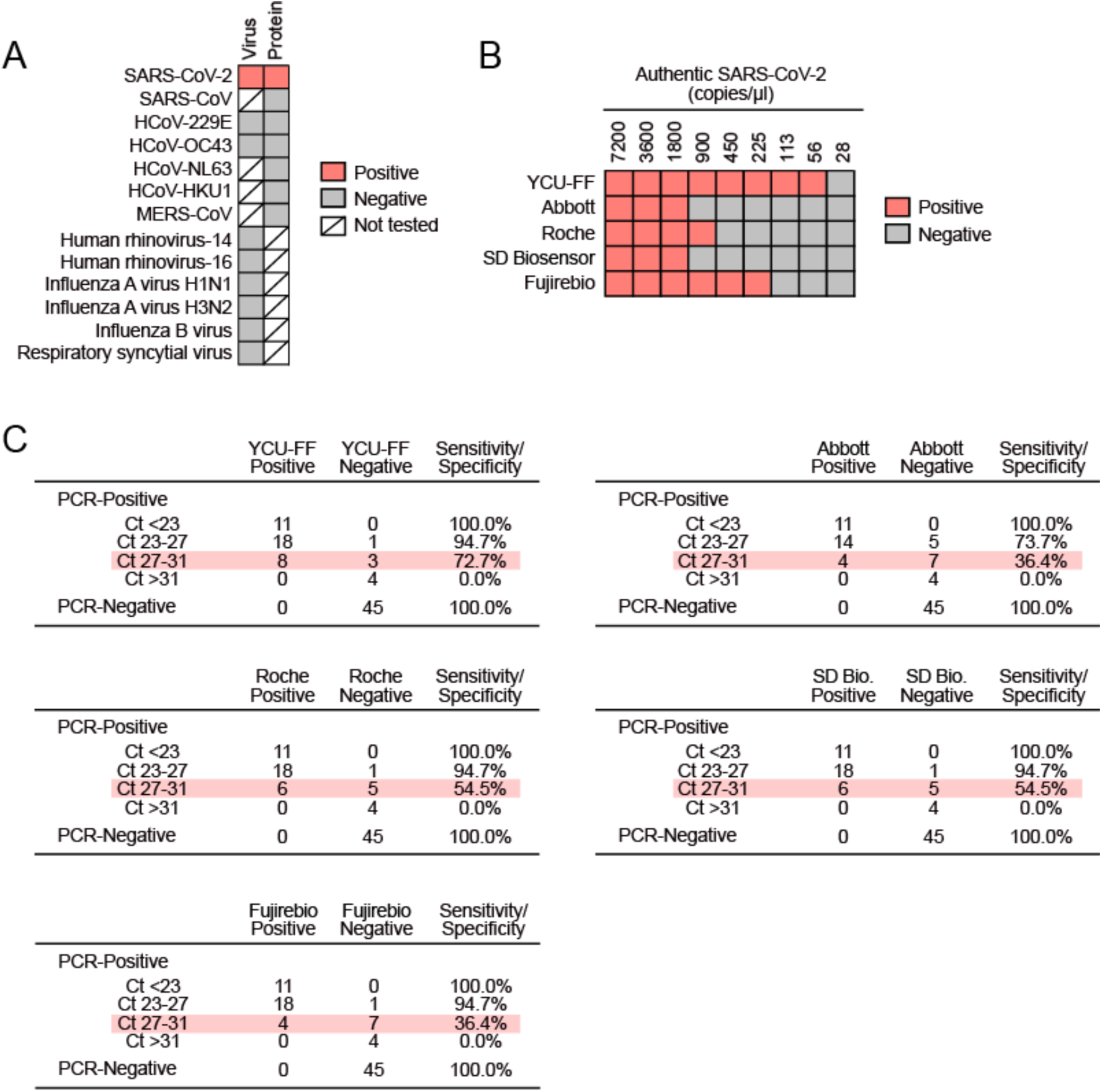
Evaluation of SARS-CoV-2 Ag-RDT. **(A)** Cross-reactivity of YCU-FF LFIA. “Virus” indicates inactivated virus (at least 10^5^ copies/µl for HCoV-229E and HCoV-OC43 or at least 10^5^ TCID_50_/ml for other viruses). “Protein” indicates recombinant NP antigen of corresponding virus (at least 200 ng/ml). **(B)** Detection of authentic SARS-CoV-2 by indicated Ag-RDTs. Graph shows the viral load, determined by RT-qPCR. Red and gray squares below indicate tested positive and negative of each Ag-RDTs, respectively. **(C)** Sensitivity and specificity of indicated SARS-CoV-2 Ag-RDTs in PCR-positive (n=45) and -negative (n=45) specimens in nasopharyngeal swabs.

We next evaluated the performance of the YCU-FF LFIA using clinical specimens (nasopharyngeal swab) that had been diagnosed by RT-PCR. With the correlation test (n=108 specimens) we observed the positive percent agreement (PPA) of the test results between the YCU-FF LFIA and the RT-PCR method to be 82.2% and the negative percent agreement (NPA) was 100% (Supplementary Figure S2). Comparison experiments using nasopharyngeal swab specimens (n=90) revealed that other Ag-RDTs did not show false positives, and none had a PPA as high as YCU-FF LFIA (Supplementary Figure S2). Notably, the YCU-FF LFIA showed superior performance compared to other Ag-RDTs in samples with moderately high Ct values (Ct = 27-31) (Figure 2C). Taken together, the YCU-FF LFIA developed in this study has definite advantages over conventional Ag-RDTs in SARS-CoV-2 detection.

## Discussion

In the ongoing fight against COVID-19, prompt detection of SARS-CoV-2 infections and timely prevention of its spread is one of the crucial strategies that can be helpful in overcoming this pandemic. Currently, NAATs are the gold standard for COVID-19 diagnosis, while quantitative antigen detection tests (such as Roche Elecsys and Abott Achitect) are also available as highly sensitive virus detection methods. However, these are not suitable for POCT. In this study, we developed the YCU-FF LFIA, a novel Ag-RDT using high quality mAbs based on the LFIA platform with output enhanced by silver halide photography technology. Compared to the conventional LFIA based Ag-RDTs currently available in the market, the YCU-FF LFIA could detect the lower concentrations of SARS-CoV-2 without the need for specialized equipment, making it a superior POCT option for rapid diagnosis of COVID-19.

The performance of antigen-capture immunoassays such as LFIA and ELISA depends on the antigen-binding specificity and optimal combination of capture and labeled antibodies used (Wada et al., 2011). Both of the mAbs used in this study had strong and specific binding to the antigen, and their epitopes were located far apart on the NP antigen surface and had no spatial interference. The binding of antibody to the antigen can be hindered if the antigenic epitope sequence gets a mutation, thereby compromising the detection sensitivity of the LFIA. Comprehensive genome analysis of 61,485 different sequences of SARS-CoV-2 NP genes revealed 1034 unique nucleotide mutations, with the most common substitution being R203K (68.09 %), followed by G204R (67.94 %) (Rahman et al., 2020). However, the none of these mutations were found in the binding region on NP of the two mAbs used in this study, implying that the YCU-FF LFIA can identify all the currently identified circulating mutants of SARS-CoV-2.

Recent studies suggest that Ag-RDT is effective around the time of onset of symptoms and a few days after when the virus is shed in sufficient amounts from the upper airway (Dinnes et al., 2020; Lambert-Niclot et al., 2020). WHO has reported that many Ag-RDT kits are to likely perform well in patients with high viral load (Ct values below 25 or above 10^6^ copies/ml). While all the Ag-RDTs tested in this study showed good concordance to high viral load samples, YCU-FF LFIA demonstrated superiority over the others with a higher probability of detection in samples with moderately high Ct values (Ct = 27-31). Similar to other commercially available kits, all PCR positive samples with low viral load (Ct above 31) were flagged negative by the YCU-FF LFIA. This presumably because of the lower amount of viral antigen or the presence of persistent, non-transmissible viral RNA. Indeed, viable virus could not be isolated at Ct values above 30 suggesting that Ag-RDTs could actually differentiate the acute transmitters from the convalescent non-transmitters which could not be distinguished by NAAT (Jefferson et al., 2020). Thus, YCU-FF LFIA may be useful for detecting individuals with infectious virus who pose the maximum risk for spread of the pandemic.

The main advantages of Ag-RDTs are rapidity, ease of use, simplicity of interpretation, no need for technical capabilities or special infrastructure and decentralized POCT capability. However, low sensitivity can hinder the detection of lower viral loads leading to poor visual output causing false negatives. YCU-FF LFIA overcomes the decrease in visibility to give results of improved sensitivity. By possessing definitive advantages over other Ag-RDTs, YCU-FF LFIA and be a better option to supplement NAATs in the diagnosis of COVID-19.

## Materials and Methods

### Virus isolates

SARS-CoV-2 strain TY-WK-521 (Matsuyama et al., 2020) was obtained from National Institute of Infectious Diseases, Japan. All experiments involving SARS-CoV-2 were performed at the biosafety level 3 laboratory at Yokohama City University School of Medicine. Human coronavirus OC43 (ATCC VR-759) and 229E (ATCC VR-740) were obtained from ATCC. Influenza viruses were cultured as previously described (Wada et al., 2011).

### Clinical specimens

Nasopharyngeal swab specimens from individuals confirmed with SARS-CoV-2 infection by RT-qPCR with N2 primer/probe set targeting the N gene were used for this evaluation (Shirato et al., 2020). Samples were stored in viral transport media at –80 °C until use. Virus load in samples was estimated from cycle threshold (Ct) value. The study protocol was approved by the IRB of Yokohama City University (B200800106).

### Development of YCU-FF LFIA

Generation of specific monoclonal antibodies against SARS-CoV-2 NP was described previously (Yamaoka et al., 2020). YCU-FF LFIA was developed over the LFIA designed in previous study (Broger et al., 2019; Wada et al., 2011). Briefly, purified monoclonal antibodies and anti-mouse IgG (Jackson Immuno Research) were diluted in 50mM Tris buffer) and dropped onto nitrocellulose membrane (ADVANTEC) to give the test and control line, respectively. The membrane was then dried at 40 °C for 30 min to immobilize antibodies. A reagent pad including monoclonal antibodies conjugated with colloidal gold (British Biocell Intonational) was placed upstream of test lines as sample application point. A test strip and two sealed pots containing a reducing reagent (containing 0.47 mol/l Fe(NH_4_)_2_(SO_4_)_2_) and a silver-ion reagent (0.31 mol/l AgNO_3_) were placed in a cartridge (Supplementary Figure S1).

### Comparison of Ag-RDTs

Commercially available Ag-RDTs (Abbott Panbio COVID-19 Ag Rapid Test, Roche SARS-CoV-2 Rapid Antigen Test, SD Biosensor Standard Q COVID-19 Ag, and Fujirebio Espline SARS-CoV-2) were used according to the manufacturers’ instructions. Test line interpretations were made by at least two people.

## Data Availability

All relevant data are available from the authors on request.

## Acknowledgements

We thank Natsumi Takaira, Kenji Yoshihara, and Kazuo Horikawa for their technical assistance.

## Author contributions

KM designed the research, analyzed the data, and wrote the manuscript; RF and YY performed the research and analyzed the data; SSJ analyzed the data and wrote the manuscript; JK, AW, and TT contributed reagents and analyzed the data; KS, HO, CK, SU, NT and EY contributed reagents; HS, NO and HH analyzed the data; AR directed the research, analyzed the data, and wrote the manuscript.

---

**Supplementary Figure 1.**
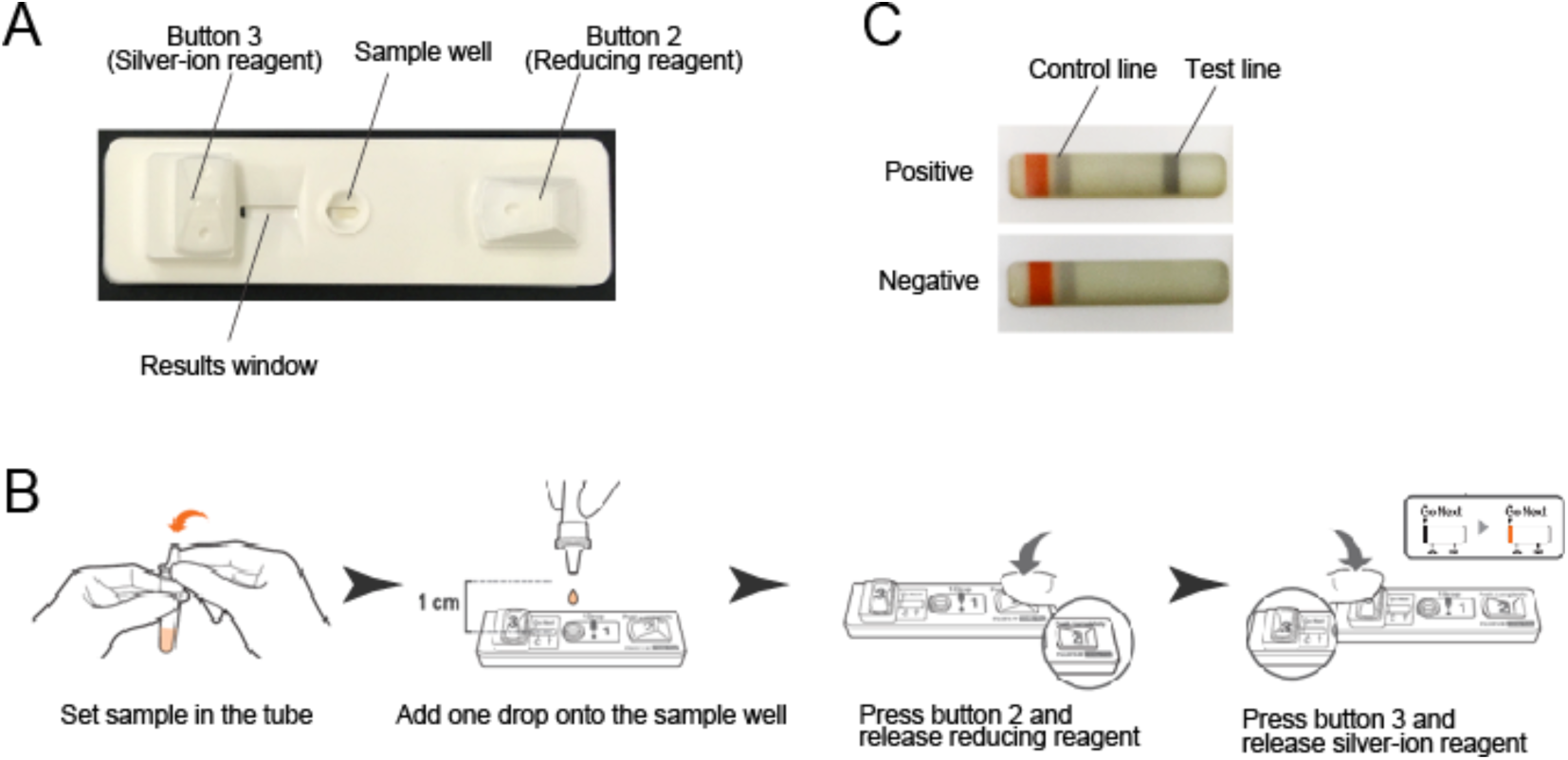
Development of YCU-FF LFIA. **(A-C)** Image of YCU-FF LFIA Cartridge (A). Samples were diluted with an extraction buffer (Tris buffer containing non-ionic surfactant) in the tube and one drop of the sample was added onto the sample well of YCU-FF LFIA cartridge. Following this, button 2 was immediately pressed to release a reducing reagent for silver amplification. After the color indicator mark turned orange (about 10 min), button 3 was pressed to release a silver-ion reagent to activate the silver amplification reaction (B). The reaction was complete immediately and the appearance of a black test line of the results window indicated a positive result, and the absence of a black test line indicated negative result (C).

**Supplementary Figure 2.**
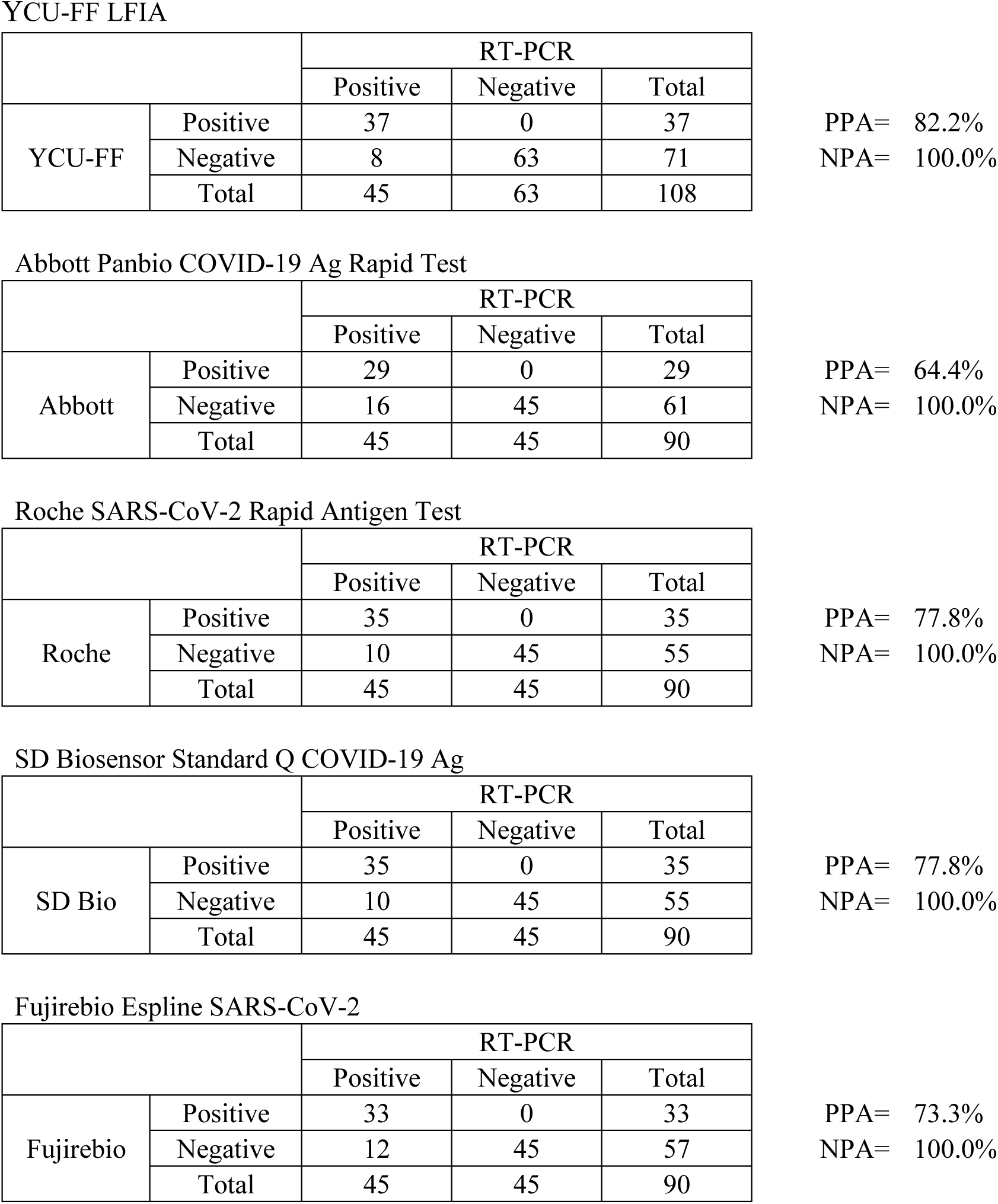
Comparison of SARS-CoV-2 Ag-RDTs. Positive percent agreement (PPA) and negative percent agreement (NPA) of the test results between the RT-PCR method and indicated SARS-CoV-2 Ag-RDTs.

